# A Drug-Specific, Half-Life-Adjusted Framework for Classifying CNS-Active Systemic Therapy Exposure During and After Radiotherapy

**DOI:** 10.64898/2026.06.11.26354463

**Authors:** Lucas Pari Mitre, Benjamin Drapkin, Michael Dohopolski

## Abstract

Clinical oncology datasets often store systemic therapy as a regimen label with a start date and an end date. Those records are clinically recognizable but can be analytically incomplete when the research question concerns whether a patient was exposed to a concurrent CNS-active drug (`cCNS-aD`) or an adjuvant CNS-active drug (`aCNS-aD`) around radiotherapy. Contemporary CNS-oncology studies usually define CNS activity by empiric drug lists and define concurrency by fixed calendar windows, although the literature shows substantial heterogeneity across both concepts. This paper proposes a generalizable framework for converting raw systemic therapy records into reproducible `cCNS-aD` and `aCNS-aD` variables, useful in subgrouping for clinical studies. The framework uses a transparent CNS scoring model based on three clinical evidence components: intracranial objective response rate, consensus CNS endorsement, and intrathecal route of administration. It then defines a pharmacokinetic exposure proxy as the recorded end date plus five half-lives plus a drug-specific steady-state accumulation term, to account for repeated dosing. Concurrent exposure is classified by overlap with the radiotherapy interval, while post-radiotherapy exposure is classified by overlap with a prespecified post-RT attribution window. The framework separately identifies post-RT pharmacokinetic persistence and post-RT treatment initiation, allowing investigators to distinguish continued exposure from true adjuvant initiation. This is a methodological framework and reference implementation. Implementation audits and endpoint-specific sensitivity analyses remain necessary before use as a definitive exposure classifier.

## 1 Introduction

The literature on systemic therapy exposure in CNS malignancies and brain metastases remains conceptually rich but can sometimes be operationally inconsistent. Naturally, most clinical datasets do not begin with analysis-ready variables such as concurrent CNS-active drug exposure and need parsing, as they derive from data extractions for medical records. Instead, they begin with a regimen name, a start date, and an end date. The analytic task is translating what was prescribed into a reproducible statement on exposure during and after radiotherapy. Concerns regarding the replicability of such translation exist, hampering reproducibility in clinical studies.

Current CNS-oncology studies generally classify a drug as CNS-active by relying on published evidence of intracranial efficacy, guideline-based drug lists, or expert judgment rather than a single universal quantitative threshold [1–4]. This practice is clinically understandable and defensible. However, this can be methodologically fragile in the setting of evolving knowledge. Two studies may both report subgroup analyses by “concurrent CNS-active systemic therapy” while using different drug lists, different timing rules, and different assumptions about whether continued biologic exposure after the nominal stop date still counts as concurrent treatment.

Timing definitions are equally heterogeneous. In glioma studies, the words “concurrent” and “adjuvant” often correspond to protocol phases, such as treatment given during radiotherapy followed by treatment resumed after radiotherapy [5–8]. In brain metastasis studies, by contrast, concurrent therapy may mean same-day treatment, treatment within 7 days, within 14 days, within 30 days, or within 90 days of radiotherapy [2, 3, 9–14]. These practical rules are chronological rather than pharmacologic. They do not distinguish between a short half-life agent whose biologic activity disappears soon after the recorded stop date and a monoclonal antibody whose meaningful exposure may persist for weeks.

Only a minority of studies explicitly acknowledge drug half-life when defining concurrent exposure [3]. Pharmacokinetic studies have long emphasized that CNS exposure is influenced by how long clinically relevant drug levels are likely to persist in the central nervous system and systemic circulation [1, 15–17]. A useful framework for `cCNS-aD` and `aCNS-aD` classification should therefore define CNS activity explicitly, specify the evidence components that generate that activity label, define the exposure window quantitatively, and define the relation between that exposure window and radiotherapy with transparent rules and equations.

This paper presents this novel framework. Its aim is to provide a rigorous and auditable method for transforming raw systemic-therapy records into analysis-ready variables that can be used across retrospective datasets, subgroup analyses, and observational outcome studies.

## 2 Framework Overview

The framework converts raw systemic therapy records into two binary exposure variables: concurrent CNS-active drug exposure (`cCNS-aD`) and adjuvant CNS-active drug exposure (`aCNS-aD`). It proceeds in five sequential steps: normalization of drug names and decomposition of combination regimens into component agents; lookup of each agent in a prespecified drug dictionary to retrieve a CNS activity class and a half-life and a steady-state accumulation factor; computation of an pharmacokinetic exposure proxy window extending the recorded treatment end date by five half-lives plus a drug-specific accumulation correction; application of overlap-based timing rules that classify each agent as concurrent, adjuvant, both, or neither relative to the radiotherapy interval; and aggregation of agent-level classifications to the patient, lesion, or treatment-course level as required by the analytic design. The drug dictionary is a reference versioned document whose CNS activity scores derive from three explicit evidence components, namely published intracranial objective response rate, consensus CNS endorsement, and intrathecal route, rather than from an opaque agent list. All steps are governed by rules declared before analysis and not modified post hoc. Figure 1 shows the full pipeline schematically.

**Figure 1.**
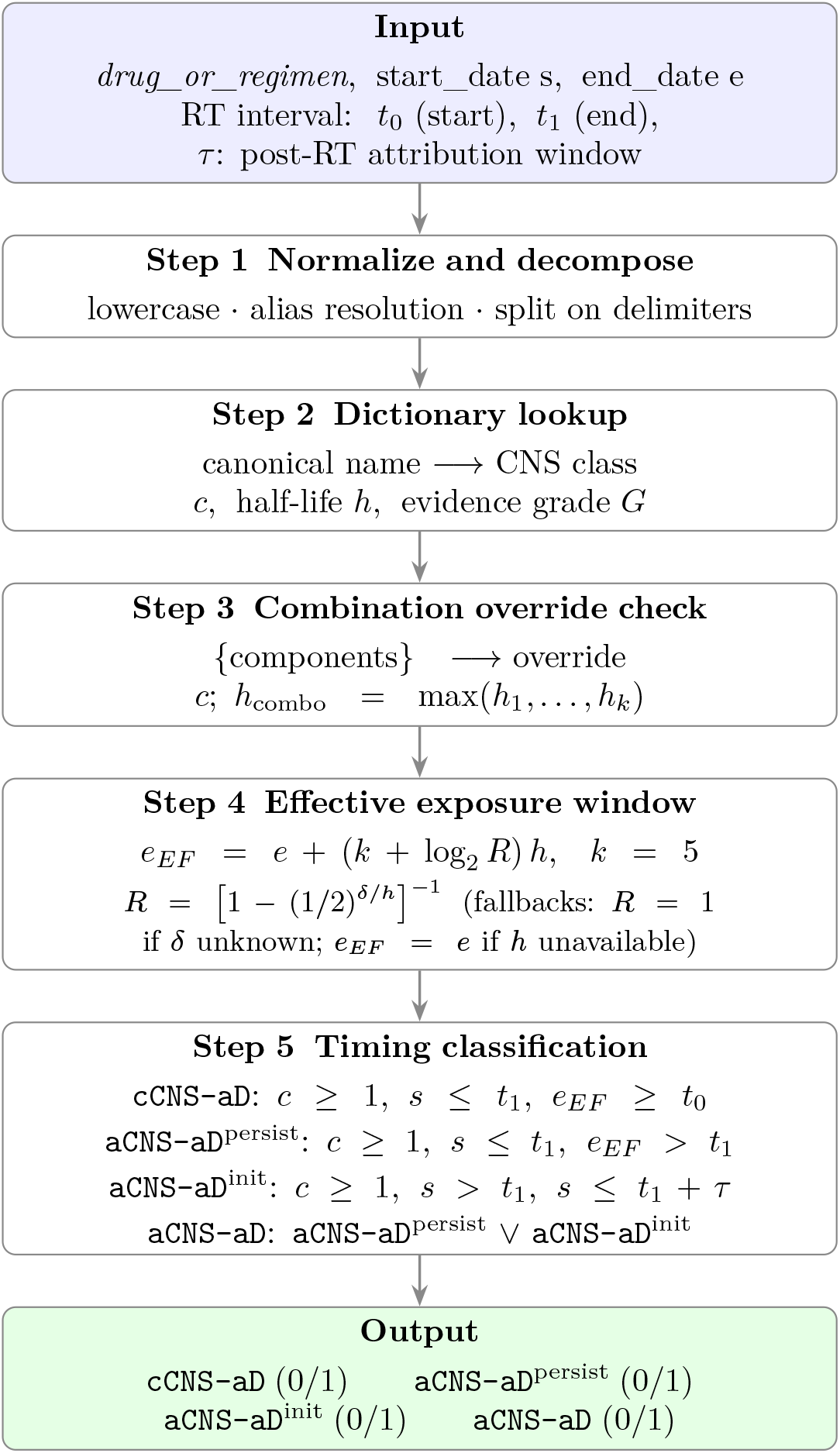
Five-step classification pipeline for deriving concurrent (`cCNS-aD`) and adjuvant (`aCNS-aD`) CNS-active drug exposure variables from raw systemic therapy records. Steps 2 and 3 together implement the drug-specific scoring and combination override logic described in Sections 5 and 8. Step 4 replaces the recorded end date with a pharmacokinetically informed effective window end that additionally accounts for steady-state accumulation under repeated dosing. Step 5 applies overlap-based timing rules rather than fixed calendar cutoffs. See Algorithm 1 for the complete procedural specification.

## 3 Required Inputs and Data Model

The framework is intended for clinical datasets in which systemic treatment is recorded as one or more drug or regimen entries, each with a start date and an end date, and in which a radiotherapy interval can be identified. The same logic can be applied at the patient level, lesion level, or treatment-course level.

Let *t*_0_ denote the first day of radiotherapy and let *t*_1_ denote the last day of radiotherapy. For a given systemic agent, let *s* denote the observed start date and let *e* denote the observed end date. Let *h* denote the drug half-life in days, derived from a predefined drug dictionary. Let *δ* denote the drug’s standard dosing interval in days and let *R* denote its steady-state accumulation factor, both retrieved from the same dictionary. Let *k* denote the prespecified half-life multiplier, fixed at *k* = 5 throughout this paper.Let *c* denote the CNS activity class assigned to that drug in the same dictionary. Let *e*_*EF*_ denote the end of the effective exposure window. Let *τ* be a prespecified post-radiation attribution window (after *t*_1_), such as 30, 60, 90 or 180 days, depending on the research question. Throughout this paper, `cCNS-aD` denotes concurrent CNS-active drug exposure and aCNS-aD denotes adjuvant CNS-active drug exposure.

## 4 Systemic Therapy Normalization

Systemic therapy is represented in the clinical source data as one or more entries of the form “drug or regimen, start date, end date.” A preprocessing step first normalizes text, date formats, and spelling variants or commercial names so that synonymous terms can map to a single standardized drug identity. Combination regimens are then decomposed into individual component agents because CNS activity and half-life are drug-specific rather than regimen-specific properties. A regimen may contain one CNS-active targeted agent and one non-CNS-active cytotoxic backbone, and collapsing the entire regimen into a single unanalyzed label would impede that distinction.

## 5 CNS Activity Dictionary and Evidence Scoring

Each normalized agent is then matched to a reference dictionary with five required parameters: a standardized drug name, a CNS activity class, a half-life measured in days, a standard dosing interval in days, and a steady-state accumulation factor carrying an explicit confidence level. The framework assumes that the dictionary is declared before analysis and is not tailored post hoc to outcome results, as shown in the worked examples below. This preserves reproducibility and aligns with the literature’s prevailing reliance on predefined evidence-aware drug lists [1–4]. The key refinement is that the CNS activity score is computed from explicit clinical evidence components.

The implemented scoring rubric is purely clinical and deliberately excludes non-outcome mechanistic features such as cerebrospinal fluid to plasma ratios, blood-brain barrier design heuristics, or other pharmacologic proxies when they are not directly tied to intracranial efficacy. The total CNS evidence score is defined as

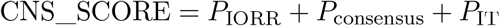

where *P*_IORR_ is the point contribution from published intracranial objective response rate (IORR), *P*_consensus_ is the point contribution from consensus CNS endorsement, and *P*_IT_ is the point contribution from intrathecal administration.

The intracranial response component is weighted most heavily because it is the most direct clinical evidence of CNS activity. If the strongest available peer-reviewed evidence, selected under the prespecified evidence hierarchy, reports an IORR of at least 60%, then

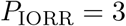

If the published IORR is 40% to 59%, then

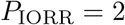

If the published IORR is 20% to 39%, then

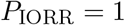

Otherwise,

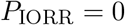

Published IORR is the closest available clinical approximation to demonstrated CNS antitumor activity, so stronger intracranial response rates deserve greater weight. The dictionary assigns IORR scores using the strongest available peer-reviewed clinical intracranial response evidence identified before dictionary lock. Prospective CNS-specific trials and prespecified CNS subgroup analyses are prioritized when available. Retrospective cohorts with defined intracranial response criteria may be used when prospective data are unavailable, provided that the evidence source, response criteria, denominator, disease setting, prior CNS therapy status, and evidence grade are recorded in the dictionary. This avoids systematically excluding agents for which prospective brain-metastasis trials were never conducted but clinically relevant intracranial activity has nonetheless been observed. When multiple studies are available, dictionary curators should avoid selectively using the most favorable estimate without context. The selected IORR should be justified according to prespecified evidence-review rules, and uncertainty should be reflected through the evidence grade G and through sensitivity analyses using stricter CNS activity thresholds.

This perspective is explicitly supported by the RANO-BM clinical trial design guideline, which recommends that for drugs with unknown CNS activity, the primary endpoint should be determining IORR and that CNS efficacy measures should also be captured bicompartmentally [1]. The guideline also cautions that although radiological IORR might be encouraging, the conclusion that a drug has CNS activity should not be defined without additional data [1], underscoring that IORR is necessary but not always sufficient on its own.

Additional contemporary evidence supports IORR’s central role while illustrating important qualifications. A 2025 analysis of CheckMate 204 found that intracranial response by mRECIST and volumetric criteria correlated with intracranial progression-free survival and overall survival, but that choice of response criteria materially affected measured rates [18]. The TRICOTEL trial used intracranial objective response rate as the primary endpoint to establish CNS activity for an atezolizumab plus BRAF/MEK regimen, and cross-study IORR comparisons were used to benchmark efficacy [19]. A 2025 ASCO meta-analysis pooling 32 trials (n=1,224) likewise used pooled CNS-ORR as the principal measure of CNS efficacy for immune checkpoint inhibitors, reinforcing IORR as the field’s default clinical metric while also highlighting heterogeneity across studies [20].

The consensus endorsement component captures drugs that are recognized as CNS-relevant by major clinical frameworks even when the intracranial response literature is sparse, mixed, histology-specific, or not reported in a way that maps cleanly onto a single IORR threshold. This component is defined as

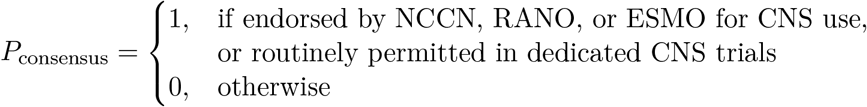

The rationale is that consensus guidance and repeated use in dedicated CNS trial settings represent clinically meaningful evidence that an agent is treated by the field as CNS-active or CNS-relevant, even when the literature does not provide a single clean intracranial response estimate. Mere allowance of patients with stable brain metastases on a systemic therapy trial is not sufficient for consensus endorsement unless the trial or guideline explicitly identifies the agent or regimen as CNS-active, CNS-directed, or clinically relevant for intracranial disease.

The intrathecal route component is defined as

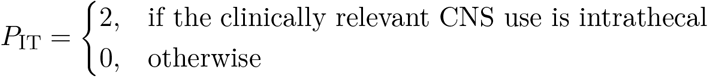

The rationale for this term is that intrathecal administration bypasses the blood-brain barrier by design. It therefore constitutes a qualitatively different basis for CNS activity than systemic penetration relevant to leptomeningeal disease, mainly. This component was introduced to prevent under-classification of agents such as methotrexate when their principal CNS efficacy derives from direct cerebrospinal fluid delivery rather than conventional systemic exposure.

The framework then maps the total score to an ordinal class *c* ∈ {0, 1, 2, 3}. Class 0 denotes no recognized CNS activity. Class 1 denotes minimal CNS activity, sufficient for analytic inclusion under this framework. Class 2 denotes moderate CNS activity. Class 3 denotes high CNS activity.

Formally,

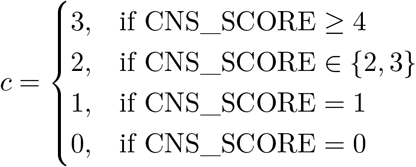

The operative exposure decision rule is

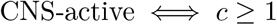

and equivalently

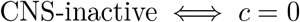

The criterion is therefore explicit. Once the agent identity is matched and the score is retrieved, any value of 1, 2, or 3 is classified as CNS-active, whereas 0 is not. This converts the hidden list-based logic used in many retrospective studies into an auditable scoring rule [1–4]. Table 1 provides representative drugs falling into each review bin under the current implementation. The full drug-by-drug scored dictionary, including component points, final class, half-life, and supporting references, is provided as Supplementary Appendix Table S1 and Supplementary File 1 (`Supplementary_Appendix_Complete.xlsx`).

**Table 1.**
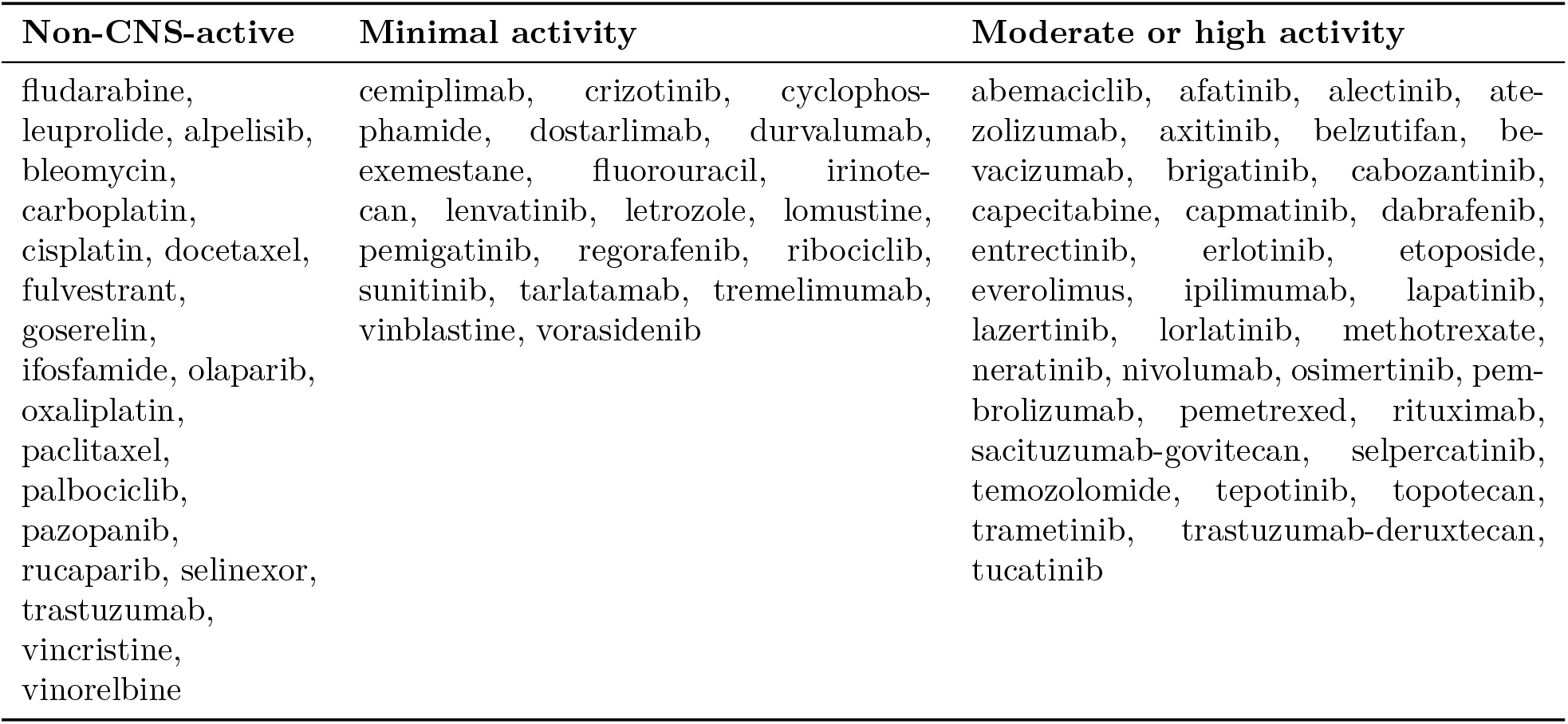
Representative example drugs by CNS activity class under the current scoring framework. Evidence grades (A–E) for each drug are provided in Supplementary Table S1.

| Non-CNS-active | Minimal activity | Moderate or high activity |
| --- | --- | --- |
| fludarabine,<br>leuprolide, alpelisib,<br>bleomycin,<br>carboplatin,<br>cisplatin, docetaxel,<br>fulvestrant,<br>goserelin,<br>ifosfamide, olaparib,<br>oxaliplatin,<br>paclitaxel,<br>palbociclib,<br>pazopanib,<br>rucaparib, selinexor,<br>trastuzumab,<br>vincristine,<br>vinorelbine | cemiplimab, crizotinib, cyclophosphamide, dostarlimab, durvalumab, exemestane, fluorouracil, irinotecan, lenvatinib, letrozole, lomustine, pemigatinib, regorafenib, ribociclib, sunitinib, tarlatamab, tremelimumab, vinblastine, vorasidenib | abemaciclib, afatinib, alectinib, atezolizumab, axitinib, belzutifan, bevacizumab, brigatinib, cabozantinib, capecitabine, capmatinib, dabrafenib, entrectinib, erlotinib, etoposide, everolimus, ipilimumab, lapatinib, lazertinib, lorlatinib, methotrexate, neratinib, nivolumab, osimertinib, pembrolizumab, pemetrexed, rituximab, sacituzumab-govitecan, selpercatinib, temozolomide, tepotinib, topotecan, trametinib, trastuzumab-deruxtecan, tucatinib |

In addition to the numeric class, each drug entry in the dictionary carries an evidence grade *G* that records the strongest class of published evidence supporting its CNS activity classification:

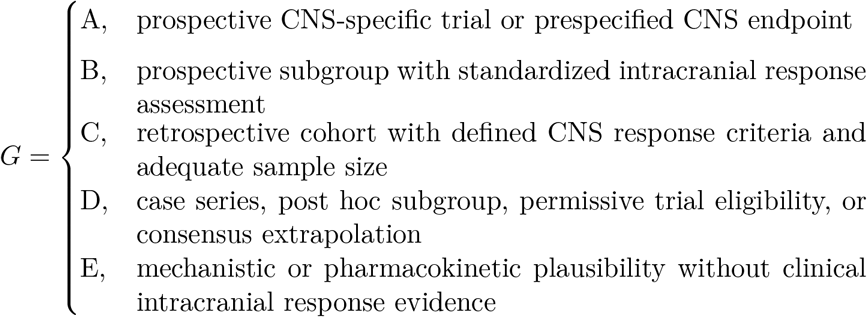

The class *c* and grade *G* are stored as separate fields in the drug dictionary. A drug may carry class 2 on a grade C evidence base, or class 1 on a grade D base; the combination conveys information that neither field alone provides. Adequate sample size is not used as a universal numeric cutoff because expected sample size differs by disease setting and drug class. The grade does not modify the class threshold or the effective-window equation.

## 6 Effective Exposure Window

The dictionary stores half-life in days and records the source used for each value. When multiple half-life estimates are available, the preferred value is the clinically relevant terminal elimination half-life from the regulatory label or a peer-reviewed pharmacokinetic study. For agents with clinically active metabolites, the dictionary may use the longer of the parent or active metabolite half-life when the metabolite is expected to contribute meaningfully to exposure. For antibodydrug conjugates, the dictionary should specify whether the intact antibody, conjugated payload, or unconjugated payload drives the exposure window. If a range is reported, the selected value and rationale should be recorded. For agents whose clinically relevant CNS use is intrathecal, the systemic elimination half-life does not govern intracranial exposure; the dictionary should instead record a route-appropriate persistence value or flag the agent so that the effective-window rule is applied on a compartment-specific basis.

The key methodological choice apart from purely chronological frameworks lies in the definition of the biologically meaningful end of exposure. For a single administration, drug decay under first-order kinetics is approximated by the remaining fraction

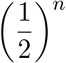

after *n* half-lives. After five half-lives, the remaining fraction is therefore

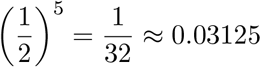

so only about 3.1% of the original drug burden remains and more than 96% has been eliminated [21–23]. The multiplier *k* = 5 is therefore used as a practical pharmacokinetic approximation for near-complete decay of a drug’s systemic presence and for a residual level that is negligible for most therapeutic purposes [21–23].

That approximation describes decay from a *single* dose. Systemic anticancer therapy is given repeatedly, and when the dosing interval is short relative to the half-life, concentration does not return to baseline between doses but climbs toward a steady-state plateau. For a drug with half-life *h* administered every *δ* days, the plateau exceeds the single-dose peak by the accumulation factor

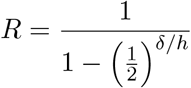

[24]. After the final dose, decay therefore begins from *R D* rather than from *D*, and the time required to reach the same residual fraction is longer by exactly log_2_ *R* half-lives. The effective exposure window end is accordingly defined as

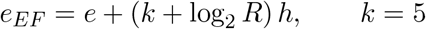

The correction log_2_(*R*) *h* does not depend on *k*. Accumulation is therefore an axis orthogonal to the choice of multiplier rather than a rescaling of it, and the two can be varied independently in sensitivity analysis.

Because a finite course cannot reach the infinite-series plateau, *R* is capped at the partial sum actually attainable in *n* administrations:

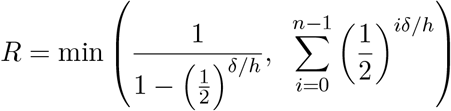

This prevents a two-dose course from inheriting the steady-state tail of a year-long one.

The magnitude of the correction is drug-dependent, and it is largest precisely where fixed calendar windows already perform worst. Agents whose dosing interval is short relative to their half-life accumulate substantially: a monoclonal antibody dosed every three weeks with a half-life of three to four weeks gains roughly one additional half-life of persistence, and agents dosed more frequently relative to their half-life gain more than two. Oral agents with half-lives under one day accumulate in relative terms but gain only hours to days in absolute terms. For agents with no meaningful accumulation the correction vanishes and the equation reduces to the single-dose form. *R* is a property of the regimen than of the individual patient, so a per-agent value derived from observed administration schedules generalizes to patients whose schedules were not directly reconstructed. Because a single observed course can move an agent’s tail by weeks, the dictionary should record the number of courses on which *R* was estimated together with an explicit confidence level, and agents whose *R* rests on only one or two observations should be inspected before use.

If the dosing interval is unknown or no administration schedule can be reconstructed, the conservative fallback is

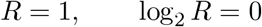

which reduces the equation exactly to the single-dose form *e*_*EF*_ = *e* + *kh*. If half-life data are unavailable altogether, the fallback is

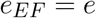

which prevents ungrounded extension of exposure beyond the recorded treatment interval. In this framework, the relevant question is not when a therapy was last charted, but how long its exposure proxy effect is plausibly present after administration stops. The rule is used to approximate pharmacokinetic persistence after the recorded end of administration. It should not be interpreted as a direct measurement of intracranial drug concentration, target engagement, radiosensitization, immune activation, or therapeutic effect.

## 7 Timing Classification

Once CNS activity and effective exposure have been defined, timing classification becomes a set of interval relations. A therapy is considered unrelated to the radiotherapy episode if its effective exposure ends before radiotherapy begins:

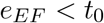

In that circumstance, the drug does not contribute to either cCNS-aD or aCNS-aD for the radiotherapy course under study.

A therapy is classified as concurrent CNS-active exposure when two conditions are both satisfied. First, the agent must be CNS-active, meaning *c*≥ 1. Second, the effective exposure interval must overlap the radiotherapy interval:

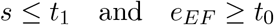

Thus,

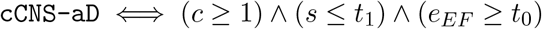

This rule is explicitly overlap-based. It does not require the drug to start during radiotherapy. A therapy that begins before radiotherapy can still be concurrent if its effective exposure persists into the radiotherapy window. This is a major advantage over study-specific calendar cutoffs such as 7, 14, or 30 days, which treat all agents as if they had equivalent biologic persistence [2, 3, 9–14].

A therapy contributes to adjuvant CNS-active exposure when the agent is CNS-active and its effective exposure proxy falls within a prespecified post-radiotherapy attribution window (*t*_1_, *t*_1_+*τ* ], where *τ* is chosen *a priori* based on the research question, such as 30, 60, 90, or 180 days. Anchoring to *τ* prevents temporally remote systemic therapies from being credited to the radiotherapy course under study. This window-bounded variable, `aCNS-aD`^*τ*^, is intentionally not mutually exclusive with `cCNS-aD`: a therapy may overlap radiotherapy and also persist into the post-RT window, in which case it is correctly classified as both. When a therapy satisfies both, analysts are recommended to retain both variables rather than forcing a single label, as the timing distinction may carry independent clinical relevance. The bounded variable decomposes into two clinically distinct sub-variables, persistence and initiation.

Setting *τ* to be unbounded recovers an initiation variable that credits any post-radiotherapy initiation of a CNS-active agent to the radiotherapy course, regardless of how remote. This is a defensible choice when the analytic question concerns whether aCNS-aD was ever given rather than whether it was given in temporal proximity to radiotherapy, but it should be declared explicitly as *τ* = ∞rather than left implicit, because the bounded and unbounded variables answer different questions and can differ substantially in datasets with long follow-up.

*Post-RT persistence* captures agents that were already active during radiotherapy and whose exposure proxy continues past the final day of RT into the attribution window. Because exposure runs continuously up to *e*_*EF*_, such an agent is present throughout (*t*_1_, *t*_1_ + *τ* ] as soon as *e*_*EF*_ *> t*_1_, irrespective of how far beyond the window it ultimately decays:

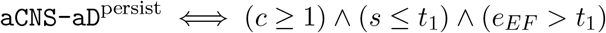

*Post-RT initiation* captures agents that begin only after radiotherapy ends but whose exposure proxy falls within the same window:

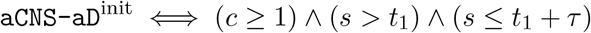

Their union recovers the bounded adjuvant set:

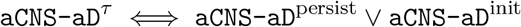

The complete classification procedure is summarized in Algorithm 1, which synthesizes the steps described in Sections 3 through 8.

### ALGORITHM 1 CNS-Active Drug Exposure Classification INPUT

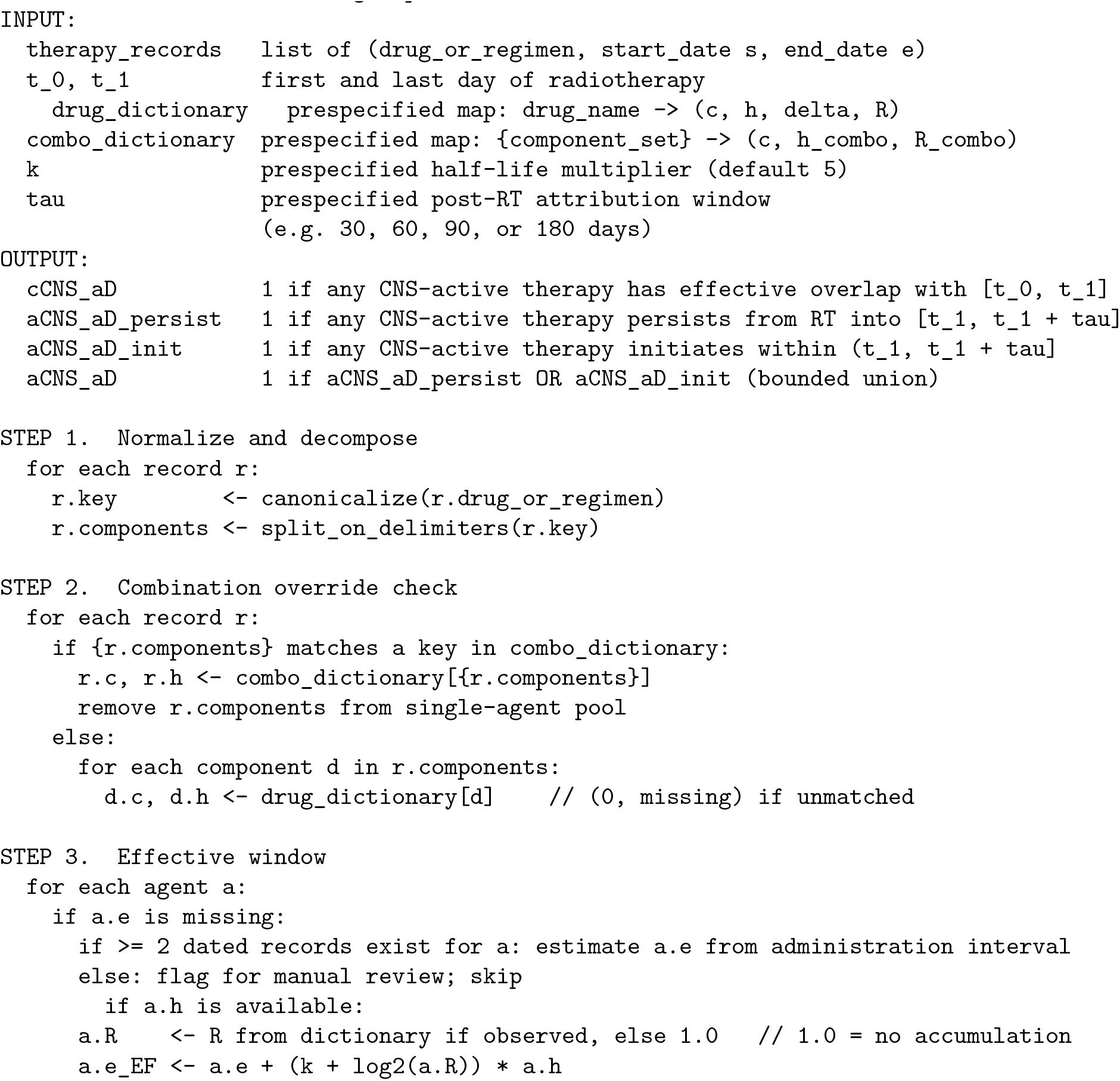
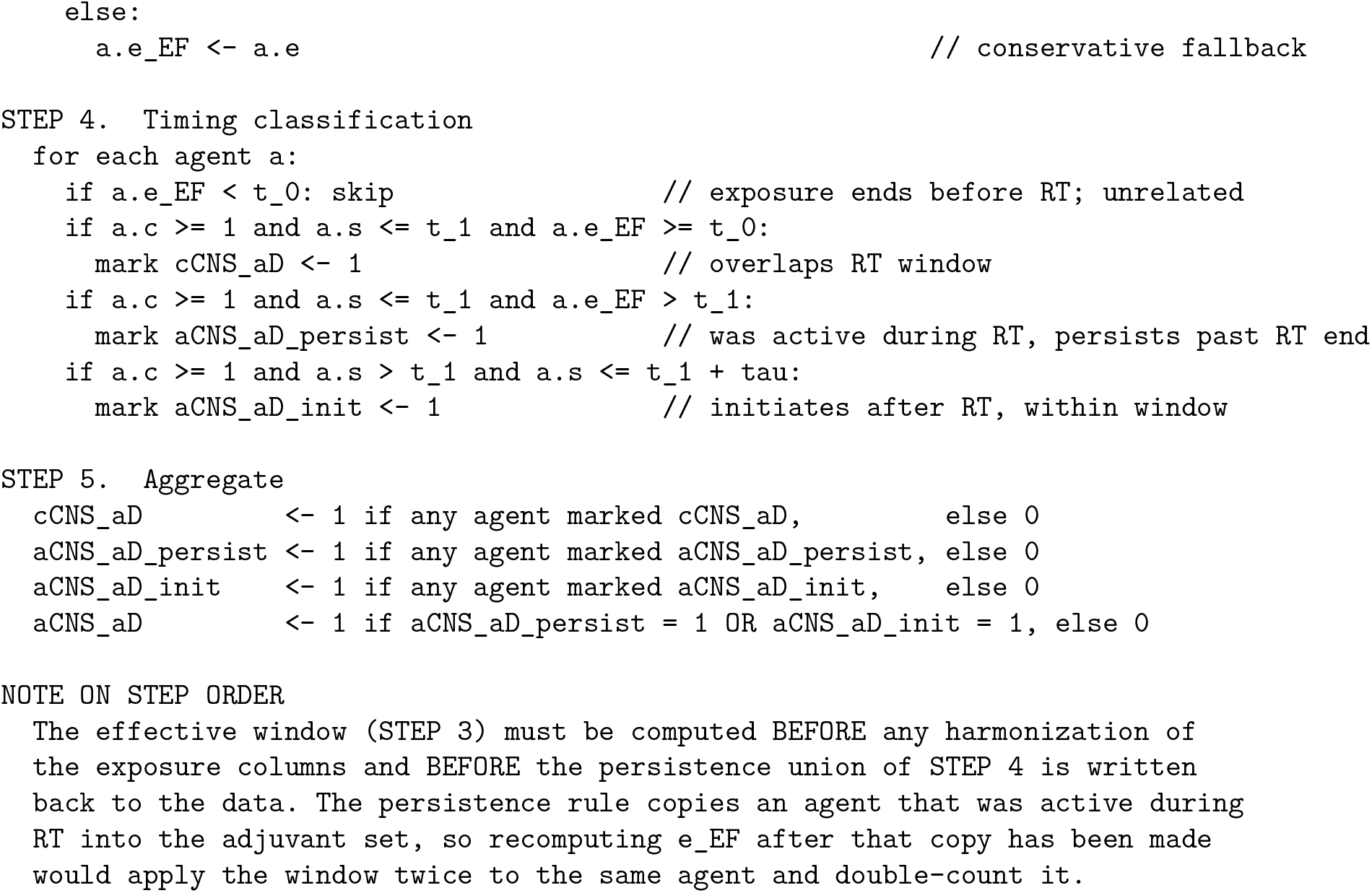

## 8 Combination-Regimen Handling

An important special case involves combination regimens whose documented intracranial activity exceeds what would be predicted from the component drugs scored independently. In that setting, the framework does not rely exclusively on the single-agent dictionary score. Instead, it applies a prespecified combination bypass. Operationally, the parser splits regimen strings on standard delimiters and normalizes common shorthand aliases to canonical component drug names; the full alias normalization table is provided in Supplementary Appendix Table S2. If all required members of a documented combination are present in the parsed regimen, the parser emits the regimen as a single combination item and removes those component drugs from further single-agent scoring. When a regimen contains a recognized combination plus additional agents, only the components included in the combination override are absorbed into the combination item; remaining agents are returned to the single-agent scoring pool. For example, tucatinib plus trastuzumab plus capecitabine given with pembrolizumab emits the HER2 triplet as one combination item and pembrolizumab as a separately scored single agent.

When a regimen contains a recognized combination plus additional agents, only the components included in the combination override are absorbed into the combination item; remaining agents are returned to the single-agent scoring pool. For example, tucatinib plus trastuzumab plus capecitabine given with pembrolizumab emits the HER2 triplet as one combination item and pembrolizumab as a separately scored single agent.

The combination item is then assigned a dedicated override CNS class rather than the arithmetic sum of the component drugs. In the current implementation, nivolumab plus ipilimumab is assigned class 3, dabrafenib plus trametinib is assigned class 3, encorafenib plus binimetinib is assigned class 3, tucatinib plus trastuzumab plus capecitabine is assigned class 2, and durvalumab plus platinum plus etoposide (carboplatin or cisplatin) is assigned class 3, with the appropriate referencing in Supplemental Table S1. The rationale is that these regimens have published brain-metastasis efficacy that is attributable to the combination itself and that would be misrepresented if the drugs were analyzed only one at a time. This bypass is therefore a regimen-level exception to the single-agent interpolation rule.

For effective-window construction, the combination item inherits a half-life equal to the maximum available half-life among its component drugs. Formally, for a regimen with component half-lives *h*_1_, *h*_2_, …, *h*_*k*_, the combo half-life is

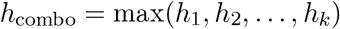

and the effective exposure end becomes

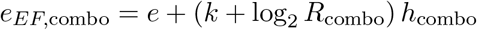

The accumulation factor requires a further rule that the half-life maximum does not. *R* depends on the ratio of dosing interval to half-life, so it must be computed per component and cannot be read off a regimen-level administration record without a compatibility check. When a combination’s components sit on different dosing scales, attributing a single reconstructed cadence to all of them produces an arbitrarily wrong result: an intravenous antibody dosed every three weeks, co-administered with oral agents taken daily, will appear to have been dosed daily and will be assigned an accumulation factor an order of magnitude too large. A regimen-level dosing interval should therefore be attributed to a component only when the components’ half-lives fall within a prespecified ratio of one another; in the reference implementation this threshold is a threefold spread, chosen so that agents on a common intravenous cycle share a cadence while an oral-plus-antibody regimen does not. Where the check fails, the components revert to the *R* = 1 fallback and the combination behaves exactly as it would under the single-dose model. Rejected attributions should be reported rather than silently discarded.

This choice intentionally prevents underestimating residual biologic exposure for combinations that include a long-persistence agent. Combination overrides should therefore be prespecified, literature-linked, and auditable. The dictionary may additionally carry an optional `half_life_driver` field for combination overrides. Its default is the maximum rule above, but the field permits an explicitly justified override when the longest half-life component is not the biologically relevant exposure driver; in that case the combination item inherits the half-life of the designated driver agent rather than the component maximum, and the rationale is recorded in the dictionary alongside its source.

## 9 Missing Data and Manual Review Rules

If the end date for a therapy record is missing, the record should not be silently assigned a concurrent or adjuvant label. When at least two dated clinical records for that drug exist in the source data, such as multiple administration or dispensing dates, the interval between them can serve as an estimated treatment duration from which a proxy end date may be derived before applying the 5*h* window. Otherwise the record should remain flagged for manual review under a prespecified missing-data rule declared before analysis.

The framework also tracks therapies that match the dictionary but remain CNS-inactive because *c* = 0, as well as unmatched therapies that fail dictionary normalization entirely. These categories should be preserved rather than discarded because they reveal whether a negative exposure call reflects true CNS inactivity or simply an unresolved mapping problem. This improves auditability and supports later refinement of the drug dictionary, and allows for real time updates given new literature findings.

## 10 Worked Examples

### 10.1 Boundary cases

Table 2 applies the classification rules to five canonical scenarios, with all dates expressed in days relative to the first day of radiotherapy (*t*_0_ = 0, *t*_1_ = 5). The scenarios are designed to show how the framework behaves at the boundaries that fixed calendar windows handle inconsistently.

**Table 2.** Boundary-case classification examples. All dates are in days relative to the first day of radiotherapy (*t*_0_ = 0, *t*_1_ = 5), with the post-RT attribution window set to *τ* = 30 days (i.e. (5, 35]). The adjuvant column is decomposed into its persistence and initiation sub-variables; aCNS-aD is their union. A dash indicates a missing value. All boundary cases assume no observed accumulation (*R* = 1, log_2_ *R* = 0), so *e*_*EF*_ = *e* + 5*h* throughout this table; the accumulation term is illustrated separately in the clinical example below.

| Scenario | $s$ | $e$ | $h$ | $c$ | $e_{EF}$ | cCNS-aD | persist | init | aCNS-aD |
| --- | --- | --- | --- | --- | --- | --- | --- | --- | --- |
| Short $h$, stopped before RT | -10 | -8 | 1 | 2 | -3 | No | No | No | No |
| Long $h$, stopped before RT | -30 | -5 | 10 | 2 | 45 | Yes | Yes | No | Yes |
| Starts during RT, continues after | 2 | 20 | 5 | 3 | 45 | Yes | Yes | No | Yes |
| Starts after RT end, within window | 10 | 30 | 5 | 3 | 55 | No | No | Yes | Yes |
| Initiates beyond window | 50 | 70 | 5 | 3 | 95 | No | No | No | No |
| Long $h$, accumulating | -40 | -5 | 20 | 3 | 115 | Yes | Yes | No | Yes |
| Missing end date | 0 | — | 5 | 2 | — | Manual | Manual | Manual | Manual |

In the first scenario, a drug stopped eight days before radiotherapy with a one-day half-life has an effective window end of day −3, which does not reach *t*_0_. The drug is unrelated to the radiotherapy episode. In the second scenario, a drug stopped five days before radiotherapy with a ten-day half-life has an effective window end of day 45. Despite ending before radiotherapy started, the drug satisfies the concurrent condition and, because it was active during radiotherapy and its exposure proxy extends past *t*_1_, it is flagged `aCNS-aD`^persist^. Note that the earlier containment form *e*_*EF*_ ≤*t*_1_ + *τ* would have erroneously dropped this agent (45 *>* 35) despite clear exposure across the entire post-RT attribution window; the overlap-based persistence rule retains it. A 7-day or 14-day calendar window would miss this drug entirely, and a 30-day window would capture it only by coincidence for this particular half-life. The third scenario behaves similarly: an agent beginning during radiotherapy and persisting afterward is both concurrent and aCNS-aD^persist^. The fourth scenario shows that a therapy beginning after the last day of radiotherapy cannot satisfy the concurrent condition (*s* = 10 *> t*_1_ = 5) but is correctly labeled aCNS-aD^init^ because it starts within the attribution window (10 ≤ *t*_1_ + *τ* = 35). The fifth scenario makes the role of *τ* explicit: a CNS-active agent initiated 45 days after radiotherapy falls outside (5, 35] and is excluded from adjuvant attribution, preventing temporally remote therapies from being credited to this radiotherapy course.

### 10.2 Clinical example

A patient with BRAF-wild-type melanoma brain metastases undergoes single-fraction stereotactic radiosurgery on day 0. Pembrolizumab, an anti-PD-1 checkpoint inhibitor with an elimination half-life of approximately 22 days, was held 42 days before treatment per the institution’s peri-procedural policy, with a last recorded administration on day −42. Pembrolizumab is administered on a three-weekly schedule, so with *δ*≈ 21 days against *h* = 22 days it accumulates to a steady-state factor of approximately *R* = 2.1, contributing log_2_ *R* ≈1.05 additional half-lives. Applying the effective window equation gives *e*_*EF*_ = −42 + (5 + 1.05) ×22≈ 91 days, against 68 days under the single-dose form. Because *s* ≤*t*_1_ = 0 and *e*_*EF*_ = 91 ≥*t*_0_ = 0, the drug satisfies the concurrent condition: `cCNS-aD = 1.` Because *e*_*EF*_ = 91 *> t*_1_ = 0, it also satisfies the adjuvant condition: `aCNS-aD = 1.` The classification here is robust to the choice of multiplier: the drug remains concurrent at every *k* from 1 to 5, although the margin narrows from 91 days at *k* = 5 to 3 days at *k* = 1, which is precisely the sensitivity that a prespecified sweep is intended to expose. A standard 30-day calendar window would have classified this drug as not concurrent, since the last dose occurred 42 days before treatment. The framework recovers the exposure because a 30-day hold does not approximate the drug’s biological persistence, which spans approximately 110 days after the last dose.

For contrast, a second entry in the same patient’s record shows cyclophosphamide discontinued 12 days before treatment (day− 12), with a half-life of approximately 0.25 days. The effective window end is *e*_*EF*_ = −12 + (5× 0.25) = −10.75 days. Because *e*_*EF*_ *< t*_0_ = 0, the drug is unrelated to the radiotherapy episode and contributes to neither variable. A 14-day calendar window would have flagged this drug as concurrent. The framework correctly excludes it because meaningful drug levels had cleared nearly eleven days before treatment.

## 11 Implementation and Reproducibility

A reference Python implementation of the full classification pipeline is provided as Supplementary File 2 (`classify_cns_exposure.py`). The script is self-contained and requires only commonly used scientific Python libraries, including pandas and numpy. It accepts three inputs: a therapy records file with one row per drug course, the scored drug dictionary provided as Supplementary File 1, and a radiotherapy records file with one row per treatment course. It returns classified output columns for `cCNS_aD, postRT_CNS_aD_persist, postRT_CNS_aD_init`, and `postRT_CNS_aD,` along with effective window end dates, the multiplier *k*, the dosing interval *δ*, the accumulation factor *R* with its confidence level and the number of courses supporting it, *τ*, CNS activity class, evidence grade, half-life source, combination override status, rejected cadence-attribution flags, unmatched-drug flags, and manual-review flags. The combination override dictionary and alias normalization table are declared at the top of the script and correspond directly to Supplementary Tables S4 and S2 respectively.

## 12 Discussion

The main methodological strength of this framework is that it uses the same conceptual language as the literature while replacing calendar timing windows with explicit pharmacologic logic. Published studies have often defined concurrency using windows ranging from same day to 90 days [2, 3, 9–14]. Such windows are pragmatic, but they can overlook meaningful biologic differences between checkpoint inhibitors, monoclonal antibodies, small molecule inhibitors, antibody-drug conjugates, and cytotoxic drugs. By extending exposure according to (*k* + log_2_ *R*)*h*, the framework adapts timing classification to the drug itself – and to how it is actually administered – rather than forcing the drug into a one-size-fits-all calendar window. The accumulation term matters most for agents dosed frequently relative to their half-life, which is the same class of agent for which fixed calendar windows are already least defensible, so the two corrections compound rather than offset.

For example, antibody-drug conjugates illustrate a dual half-life problem: the intact construct and the cytotoxic payload may have very different persistence, so a fixed 21- to 30-day window can misrepresent when meaningful exposure is still present [25,26]. BRAF/MEK combinations illustrate a different mismatch. Dabrafenib has a short parent half-life and active metabolites with longer persistence, whereas trametinib persists substantially longer, so a short safety-based hold around radiotherapy does not imply pharmacologic absence of the regimen [27–30]. Bevacizumab illustrates a long-tail problem. It is often held around procedures for safety reasons, but its prolonged half-life means that calendar washout and biologic exposure are not equivalent concepts [31]. At the opposite extreme, continuous-infusion or very short half-life agents such as blinatumomab can lose meaningful exposure rapidly with interruption, making broad calendar categories especially crude [32]. Checkpoint inhibitor groupings can also overlook meaningful variation when agents with different persistence and limited induction schedules are treated as if they were interchangeable over a shared 30-day window [33]. These examples motivate a drug-specific framework because the error introduced by pure calendar categorization is not random; it is largest precisely for agents with unusual or clinically heterogeneous pharmacokinetics.

The framework also improves reproducibility by separating evidence from logic. The classification rules do not change from study to study; only the frozen dictionary version, half-life values, and prespecified thresholds are declared. This allows investigators to update the evidence base as new CNS efficacy data emerge while preserving an auditable record of which dictionary version was used for a given analysis.

## 13 Limitations

This framework does not solve every ambiguity in CNS drug classification. It still depends on the quality of the source regimen strings, the quality of the drug dictionary, and the validity of the evidence used to assign CNS activity scores. The choice of five half-lives is pharmacokinetically sensible, but it remains a modeling assumption rather than a direct measurement of intracranial target engagement. Blood-brain barrier heterogeneity, tumor-specific permeability, dose intensity, intermittent scheduling, and drug-drug interactions are not fully captured by a single half-life adjusted exposure equation [1, 15–17]. For these reasons, the framework should be viewed as a disciplined and transparent approximation of biologically meaningful exposure, not as a perfect surrogate for intracranial pharmacodynamics.

The framework also assumes that systemic therapies are being evaluated in the clinically appropriate tumor-type and biomarker context. For example, the CNS activity classification of a targeted therapy is intended to apply when the corresponding molecular alteration and disease setting are present. The framework does not independently adjudicate whether a drug is biologically appropriate for a given patient’s cancer. Future dictionary versions could incorporate explicit tumor-type, biomarker, CNS-compartment, prior-local-therapy, and route-specific dosing-schedule fields.

## 14 Conclusion

The conversion of raw clinical systemic therapy records into `cCNS-aD` and `aCNS-aD` is a methodological problem that deserves explicit treatment. The literature currently relies heavily on pragmatic agent lists and heterogeneous calendar windows [1–4, 9–17]. The framework presented here offers a more precise alternative. It classifies agents as CNS-active when their predefined score satisfies *c* ≥1, defines effective exposure as *e*_*EF*_ = *e* + (*k* + log_2_ *R*)*h* with *k* = 5, and classifies concurrent and adjuvant exposure by overlap with the radiotherapy interval. In doing so, it preserves clinical interpretability while replacing vague chronological proximity with explicit, auditable, pharmacokinetically informed rules.

A central practical advantage of this approach is that the drug dictionary functions as a versioned document. Because CNS activity scores and half-life values are stored as explicit, named parameters rather than embedded assumptions, incorporating new published evidence requires updating only the relevant dictionary entry, leaving the classification logic itself unchanged. This decoupling of evidence from logic allows the framework to remain current as the literature evolves without rewriting the analytic pipeline. It is important to add that, for each analysis, the dictionary must be frozen before outcome analysis and archived with a version number, date, commit hash or DOI, evidence-review date, and full change log. The master dictionary may evolve, but each study must cite the frozen analytic dictionary version.

## Supporting information

Supplementary File 2 - PyScript

## Data Availability

All data produced in the present study are available upon reasonable request to the authors

